# A clinical trial to evaluate the dayzz smartphone app on employee sleep, health, and productivity at a large US employer

**DOI:** 10.1101/2021.10.06.21264618

**Authors:** Rebecca Robbins, Matthew D. Weaver, Stuart F. Quan, Jason P. Sullivan, Mairav Cohen-Zion, Laura Glasner, Salim Qadri, Charles A. Czeisler, Laura Barger

**Affiliations:** Division of Sleep and Circadian Disorders, Departments of Medicine and Neurology, Brigham & Women’s Hospital, Boston, Massachusetts, United States of America; Division of Sleep Medicine, Harvard Medical School, Boston, Massachusetts, United States of America; dayzz Live Well Ltd, Herzliya, Israel; The Academic College of Tel Aviv-Jaffa, Tel Aviv, Israel; Sheba Medical Center, Ramat Gan, Israel

**Keywords:** Sleep, smartphone application, mobile health (mHealth), workplace wellness

## Abstract

Sleep deficiency is a hidden cost of our 24-7 society, with 70% of adults in the US admitting that they routinely obtain insufficient sleep. Further, it is estimated that 50-70 million adults in the US have a sleep disorder. Undiagnosed and untreated sleep disorders are associated with diminished health for the individual and increased costs for the employer. Research has shown that adverse impacts on employees and employers can be mitigated through sleep health education and sleep disorder screening and treatment programs. Smartphone applications (app) are increasingly commonplace and represent promising, scalable modalities for such programs. The dayzz app is a personalized sleep training program that incorporates assessment of sleep disorders and offers a personalized comprehensive sleep improvement solution. Using a sample of day workers affiliated with a large institution of higher education, we will conduct a single-site, parallel-group, randomized, waitlist control trial. Participants will be randomly assigned to either use the dayzz app throughout the study or receive the dayzz app at the end of the study. We will collect data on feasibility and acceptability of the dayzz app; employee sleep, including sleep behavioral changes, sleep duration, regularity, and quality; employee presenteeism, absenteeism, and performance; employee mood; adverse and safety outcomes; and healthcare utilization on a monthly basis throughout the study, as well as collect more granular daily data from the employee during pre-specified intervals. Our results will illuminate whether a personalized smartphone app is a viable approach for improving employee sleep, health, and productivity.

## Introduction

Sleep deficiency is a hidden cost of our 24-7 society, with 70% of Americans admitting they routinely obtain insufficient sleep.^1^ The issue is pervasive in our workforce, as 30% of United States (US) workers report sleeping less than 6 hours per night.^2^ Additionally, approximately 50-70 million individuals in the US have a sleep disorder.^3^ Undiagnosed and untreated sleep disorders are associated with short and long-term health consequences for the individual and generate substantial direct and indirect costs for the employer.

Diminished alertness as a result of sleep deficiency or undiagnosed and untreated sleep disorders are associated with increased costs to employers. Costs attributable to sleep deficiency in the US were estimated to exceed $410 billion dollars in 2015, equivalent to 2.28% of the gross domestic product.^4^ Despite the importance of healthy sleep for workplace productivity and employee health and safety, nationally representative survey data collected from employers in the US shows that fewer than 10% of worksites report offering sleep-focused programs.^5^

Sleep disorders are very common, with 30-40% of employees screening positive for at least one common sleep disorder.^6–9^ Approximately 85% of those with sleep disorders are undiagnosed and untreated.^6,10^ Screening positive for a sleep disorder has been associated with a myriad of adverse health consequences, including increased risk of cardiovascular disease, diabetes and depression.^6,10^ Sleep disorders also compromise safety, being associated with an increased risk of motor vehicle crashes^10^ and administrative errors or safety violations.^6^ Further, research has shown that hospital workers who screened positive for a sleep disorder had an 83% increased incidence of adverse safety outcomes (including medical errors, needlestick injuries, and motor vehicle crashes) over a 6-month interval.^9^

Research has shown that workplace health programs focused on employee sleep or sleep disorders screening are effective for improving employee sleep, health, well-being, and workplace outcomes. In a randomized controlled trial of an in-person sleep education and sleep disorders screening intervention, researchers found there was a 24% reduction in injuries and a 46% reduction in disability day usage over the following year.^10^ In a national trucking company, each employee diagnosed and treated for sleep apnea through an employer-sponsored program, saved nearly $3,000 per employee in health care costs annually.^11^

In part catalyzed by the COVID-19 pandemic, there has been a widespread adoption of flexible and work-from-home policies among employers,^12^ and many forecast that worksites are likely to feature these policies when employees return to work.^13^ These trends present new challenges to employee health and wellbeing when employees manage both work and personal obligations inside the home. The trend toward increased work-from-home policies and flexible work schedules may also presents opportunities for digital health programs for improving employee health. Digital platforms, such as websites, virtual coaching, and smartphone applications, may extend sleep health and wellness programs and offer tangible benefits to employers, such as scalability and cost effectiveness. These digital and mobile health (mHealth) approaches are particularly promising as they have the ability to monitor user behavior (e.g., time spent sleeping or number of steps taken) and to personalize messages tailored to a user’s lifestyle and needs, ^14,15^ aimed at nudging towards health promoting behaviors and attitudes.

The purpose of this study is to conduct a randomized controlled trial to evaluate a smartphone app designed to improve employee sleep health and improve workplace outcomes. We will ascertain the feasibility and acceptability of the smartphone app and determine whether use of the app will improve sleep, health, mood, productivity, safety, and productivity among a large employee population in the US Northeast.

## Materials and methods

### Study Design

We will conduct a single-site, randomized, two-arm, parallel-group controlled trial using a waitlist control design. We will enroll a target of 1,000 daytime employees into the study, 500 randomly assigned to the experimental group and 500 to the control group. The experimental group will receive access to the dayzz app as well as to a short sleep health and wellness (SHAW) education program, developed by the Sleep Matters Initiative which includes screening for common sleep disorders. The control group will receive the dayzz app and SHAW at the end of the study. The study recruitment was originally intended to be conducted in-person. However, due to the onset of the COVID-19 pandemic at the start of study implementation, the study recruitment, enrollment, and follow-up procedures were re-designed to be completed entirely online. Please see the Fig 1 for a flowchart of enrollment, interventions, and assessments.

**Figure 1.**
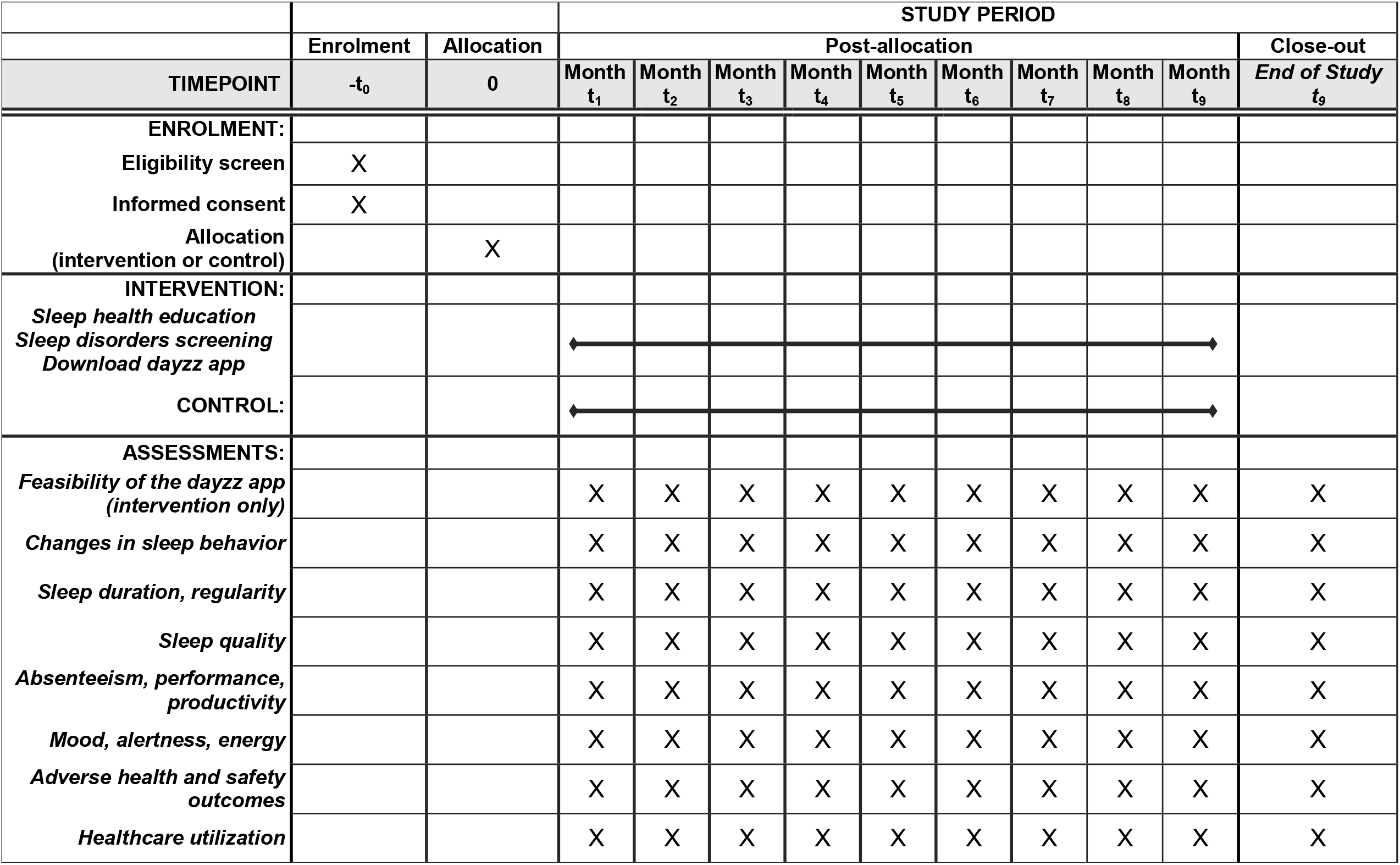
SPIRIT flowchart of the schedule of enrolment, interventions, and assessments.

### Participants

Eligible participants will be daytime workers employed by a large research and teaching organization in the US Northeast. Participants must be employed by the organization, work during the daytime, own and use a smartphone, and regularly use smartphone apps (i.e., once per week). Exclusion criteria for this study are regularly working evening, night or rotating shifts and pregnancy or breast-feeding.

### Recruitment and Retention

Recruitment will include two emails that will be sent to more than 20,000 members of the organization with an invitation to visit the study website for more information. An advertisement for the study will also be posted on a website hosted by the organization that is devoted to research and study recruitment, with a wide circulation (>10,000 visitors monthly). Finally, will post targeted social media advertisements to individuals who report the organization as their primary employer on Facebook and LinkedIn.

Potential participants will be directed to an online landing page with more information about the study. An online screener will be automatically scored and used to determine eligibility according to the study inclusion/exclusion criteria, resulting in either an invitation to provide consent and complete the baseline procedure, or a message indicating they were not eligible. Due to the virtual nature of this study, consent was collected via electronic signature. Participants were informed they could stop at any time, and who had questions were encouraged to email or call the study team. Participant responses will be de-identified and results published in aggregate. No identifiable data will be disclosed to the employer.

After providing consent via the online landing page, participants were automatically randomized by a computer program. Due to the multiple steps necessary to enroll in the experimental arm, we weighted assignment to the experimental group 1.3 to 1 to ensure equal enrollment in each group.

All enrolled participants will receive $25 for completing the baseline procedures, a t-shirt ($10 value) for completing the first monthly sleep diary, and a $20 gift card for completing the end-of-study questionnaire. Each month, participants who complete the monthly questionnaire which they receive via email will be entered in a raffle to win one of two pairs of Bluetooth wireless headphones ($250 value each). Finally, for each study component completed, participants receive chances for a raffle of gift cards, ranging in value from $100 to $500, at the end of the study. Participants who are unresponsive to emailed questionnaires will receive up to 2 reminder follow-up emails.

### Study Arms

#### Experimental arm

After consenting, participants randomized to the experimental group will take a short online initial contact questionnaire to collect basic demographics. They will then view a 20-minute sleep health education video. Immediately after the presentation, the participant will be presented with a short sleep disorders risk assessment which displays the results upon completion, either high risk for a sleep disorder (obstructive sleep apnea, insomnia, or restless legs syndrome) or low/no risk a sleep disorder. Participants are then presented with a baseline questionnaire to assess sleep, health, and lifestyle. Finally, the experimental group participants will be invited to download the study app (“dayzz”). They will be encouraged to use the dayzz app throughout the study (up to 9 months, depending on date of enrollment).

#### Control

Participants assigned to the waitlist control condition will, at baseline, complete the same initial contact and baseline questionnaires. Participants in the waitlist control group will complete the eDiary and monthly and end-of-study questionnaires. At the end of the study, waitlist control participants will receive the intervention (SHAW and dayzz app).

Each participant will also complete a daily electronic sleep diary (eDiary) sent via email for one week within the first month of enrollment and for one week within the third month of the study. For up to 9 months following the baseline procedure, on the 28th of each month, each participant will receive an email with a link to the monthly questionnaire. At the end of the study, each participant will receive a link to complete the end-of-study questionnaire. The eDiary and all questionnaires are completed online. This study was approved by the Partners HealthCare Institutional Review Board (IRB) on February 10, 2020 (Protocol #: 2019P003277). Please see the Fig 2 for a flow chart describing the study procedures. Per the IRB, a Data Safety Monitoring Board was not required for this clinical trial.

**Figure 2.**
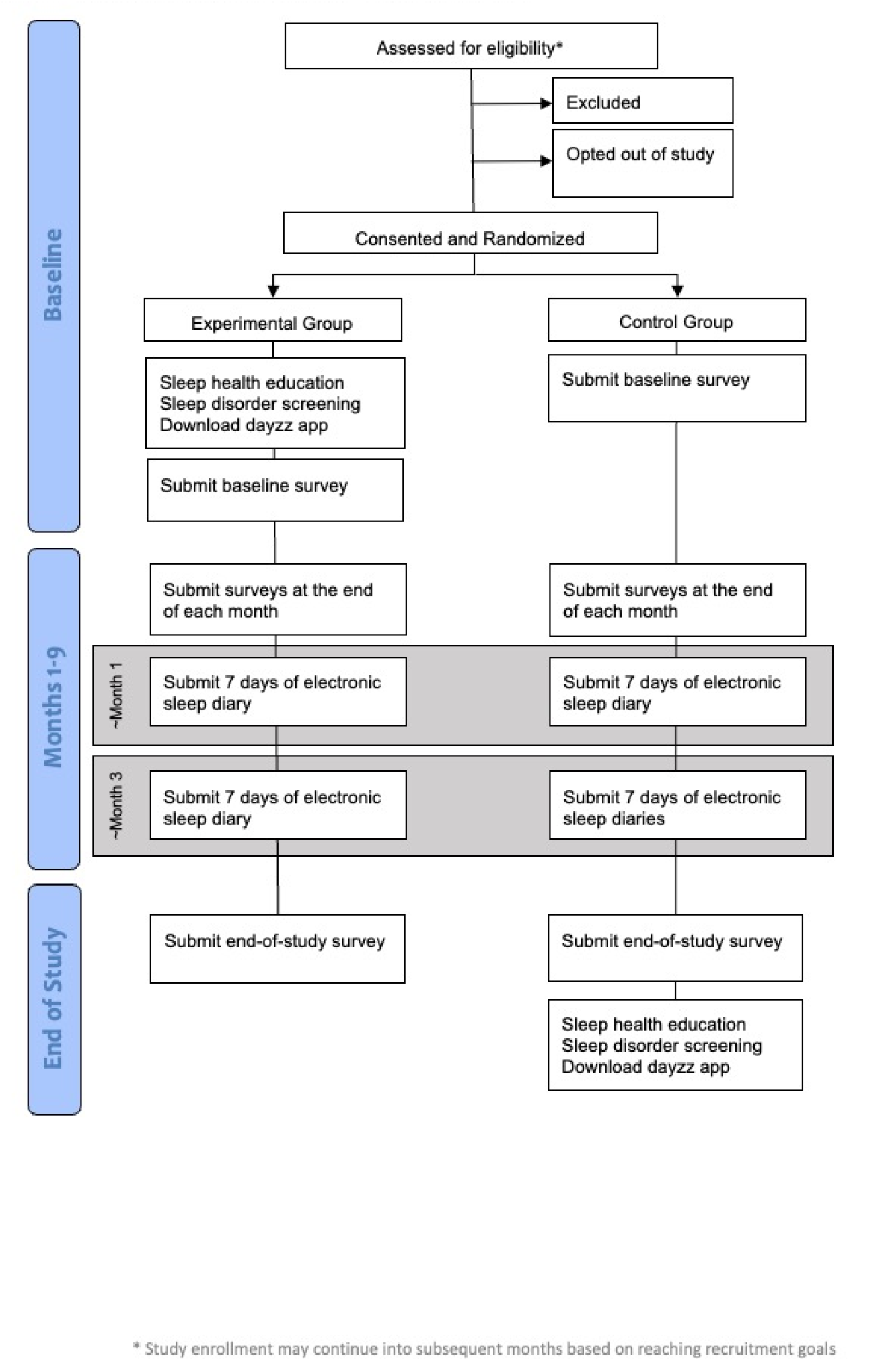
Flowchart of study procedures.

#### The dayzz App

The dayzz app is a personalized, digital sleep assessment and treatment program offered via smartphone. The dayzz app begins with a brief onboarding and registration process, followed by the administration of a clinically validated digital sleep disorders questionnaire (DSQ).^16^ The DSQ uses machine learning algorithms, previously validated against gold-standard physician sleep diagnoses, to identify people at high risk for four common sleep conditions: insomnia, delayed sleep phase syndrome (DSPS), insufficient sleep syndrome (ISS), and suspected obstructive sleep apnea. The information regarding sleep disorders is used to best tailor the sleep training plan and track progress for each individual user.

During the onboarding process, the user has the option to connect an external sleep monitor (e.g., Fitbit, Garmin, Whithings, Oura, Apple Watch) or a digital health data platform (e.g., Apple Health) and permit dayzz to stream live sleep and activity data into the app and prepopulate nightly sleep trackers. Users who do not connect wearable devices can manually enter daily sleep data using the app’s sleep tracker feature. Users can also enter sleep information retroactively and can override wearable sleep data by manually editing their sleep information within the sleep tracker feature. Users can review their sleep progress over time within the app as well, using a range of features reflecting the user’s sleep and progress during the sleep training.

Based on the user’s sleep assessment outcomes, behavioral data, and collected sleep data (subjective and/or objective), the dayzz app offers the user a personalized sleep training and improvement plan. Training plans consist of modules aimed at improving the user’s specific sleep issues. Modules are tailored to deliver evidence-based therapies for the specific sleep issue users report, for example, cognitive-behavioral skills and strategies for users at risk for insomnia. All users are provided with modules covering basic sleep hygiene principles and tools, such as bedroom environment optimization (integrating smartphone noise and light sensors), white noise audios, and written or auditory content to promote sleep health.

In addition, for users identified with suspected sleep apnea syndrome, a unique module was designed specifically for the purposes of this study, which recommends the user may benefit from a more in-depth sleep evaluation by a sleep physician. The module offers users the option to voluntarily set up visits (virtual or in-person) at a local sleep center for further evaluation (including home sleep studies per the doctor’s referral), track their progress (non-medical), and remind them of their scheduled appointments. Individuals diagnosed with Obstructive Sleep Apnea and prescribed a Continuous Positive Airway Pressure (CPAP) device by the physician, will be offered a training module to support the adjustment and adherence to their CPAP treatment.

To encourage user engagement with the dayzz app and increase adherence to intervention modules, the platform integrates contextual coaching strategies in the form of real-time intervention prompts to encourage positive sleep health-related behaviors and adherence to the training program. Such prompts include push notifications, chat messages, in-app overlays and email messages providing individual sleep feedback and task recommendations, sleep knowledge, rewards in the forms of badges for sleep-related behavioral change efforts and sleep metric achievements. The timing of the prompts is optimized for each individual user and based on smartphone sensor data, including location (GPS, WiFi connectivity), movement (accelerometer), time of day, and smartphone active/idle time. Users are asked but are not required to accept app permissions, such as location, motion, and fitness data access, to provide an enhanced user experience.

### Measures

We will describe the feasibility and acceptability of the app, and evaluate changes in sleep, health, safety, healthcare utilization, and workplace outcomes using self-reported questionnaires.

#### Feasibility and acceptability of the app

We will collect feasibility and acceptability data in the experimental group only, using two ways. First, participants in the experimental group will provide subjective ratings of acceptability of the app on the end-of-study questionnaire. Specifically, participants will be asked to respond to several questions, including “Did the study app provide helpful informationã,” “Would you recommend the app to othersã” and “Did you find the app easy to useã” Responses will be collected on scales from 1 (“Not at all helpful or “Not at all”) to 7 (“Very helpful” or “Very much so”). Second, we will collect objective app usage data provided by the developers, including: a) number of modules and tasks within each module completed on the app, per participant; b) number of features utilized, such as embedded apps (e.g. Spotify), reminders (e.g., last cup of coffee, alarm), use of favorites gallery); c) use of the chat option; d) responsiveness to notifications; f) number of achievements/badges received for reaching predefined goals; g) number of days and times the app is accessed; and h) integration of wearable devices; to characterize engagement with the intervention.

#### Changes in sleep behavior

We will evaluate changes in sleep behaviors on the monthly and the end-of-study surveys. Using a checklist, participants will be asked to select the healthy sleep changes they have made since starting the study. Specifically, participants are asked “During this study, have you changed any sleep-related behaviors to improve your sleep since participating in the study (check all that apply)ã” Participants have the option to select changes they may have made, such as “Go to bed earlier,” “Keep a more consistent sleeping schedule,” and “Set an alarm to remind you of your bedtime.” Participants are also able to select “other” and identify the healthy sleep change they made since joining the study.

#### Sleep duration and regularity

Participants will self-report their sleep duration and timing on the eDiary. The Sleep Regularity Index is the percentage probability of an individual being in the same state (asleep vs. awake) at any two time-points 24 h apart, averaged across the study.^17^ The index is scaled so that an individual who sleeps and wakes at exactly the same times each day scores 100 (better outcome), whereas an individual who sleeps and wakes at random scores 0 (worse outcome).

#### Sleep quality

Sleep quality will be measured using the Pittsburgh Sleep Quality Index (PSQI).^18^ The Pittsburgh Sleep Quality Index (PSQI) is an effective instrument used to measure the quality and patterns of sleep in adults. It differentiates “poor” from “good” sleep by measuring seven domains: subjective sleep quality, sleep latency, sleep duration, habitual sleep efficiency, sleep disturbances, use of sleep medication, and daytime dysfunction over the last month. The participant self-rates each of these seven areas of sleep. Scoring of the answers is based on a 0 to 3 scale, whereby 3 reflects the negative extreme on the Likert scale. Global scores range from 0-21 with a score of 5 or greater indicating poor sleep.

#### Workplace absenteeism, performance, and productivity

Absenteeism, performance, and productivity will be evaluated on the monthly questionnaire using the World Health Organization (WHO) Health and Work Performance Questionnaire Short Form.^19^ Participants will be asked the number of hours worked in a typical week. Unscheduled absences and disability day usage will be collected by self-report. Presenteeism questions ask the worker to compare both their performance to their best possible performance, and separately, their performance to the performance of most workers in a similar job. A ratio of workplace performance relative to ideal or possible performance will be calculated. Absenteeism and presenteeism can be converted to workplace cost by multiplying the number of hours lost by the individuals’ salary, which is reported on the demographic questionnaire.

#### Mood, alertness, and energy

Mood, alertness, and energy will be assessed on the eDiary. Specifically, participants will be asked to report, using 100 mm visual analog scales, their: 1) mood from “Sad” (0) to “Happy” (100); their 2) alertness from “Sleepy” (0) to “Alert” (100); and 3) energy from “Sluggish” (0) to “Energetic” (100). Higher scores indicate a better outcome. On the monthly questionnaires, we will also assess mood on a 7-point scale from “Very poor” (0) to “Very good” (7), alertness from “Very poor” (0) to “Very good” (7) and energy from “Very low” to “Very high” (7).

#### Adverse health and safety outcomes

Motor vehicle crashes and near-crashes will be captured via self-report on the monthly questionnaire, consistent with previous studies.^7,20^ Participants will be asked, “In the last month, did you have any motor vehicle accidents or crashes (actual collisions) in which you were driving,” and “In the last month, did you have any near miss motor vehicle accidents or crashes (narrowly avoided property damage or bodily harm) in which you were driving.” Participants who responded yes will then be asked to provide the number of times that each outcome occurred during the month. We will also assess injuries by asking participants “In the last month, how many injuries did you have” and attentional failures by asking participants the number of times they “Nod off or fall asleep during meetings at work,” “…on the telephone,” “…while driving,” and “…while stopped in traffic.”

#### Healthcare utilization

Healthcare utilization will be reported on monthly surveys. Participants will be asked to report interaction with the healthcare system, including visits to their primary care provider, emergency room, urgent care, or a specialist. They will further be asked to report any laboratory tests and diagnostic evaluations, including imaging, home sleep tests, and other examples of healthcare utilization. Healthcare costs will be estimated for each interaction through applying national average costs for each visit, test, and/or procedure.

### Power Analysis

The power analysis was performed on the anticipated cohort of 1000 active participants providing adequate data for comparison (500 in each arm of the protocol). Power was estimated for each of the aims using GPower version 3.1.9.4. We will be able to detect slightly smaller effects for continuous variables compared to binary variables if we choose to dichotomize in the analysis (e.g. any motor vehicle crash vs. none). A sample size of 500 in each group will enable us to detect an effect size of 0.16 between the groups (even if the impact is small, we will be able to detect a significant difference). This effect size roughly translates to a relative risk of 1.20. If fewer participants enroll than we expect, we will still be able to detect a small difference in effect size if we enroll at least 600 participants.

### Statistical Plan

We plan to analyze anonymized data using an intention-to-treat approach (ITT). All outcome measures will be compared by assignment to the experimental and waitlist control groups in the primary analysis. The primary ITT analysis will include all participants who are randomized in the study. Secondary analyses will compare outcomes using an as-treated approach and an adherent to treatment approach. The as-treated approach will compare those who download dayzz to those who do not, regardless of their assigned randomization. We anticipate some cross-over between groups due to sharing of the app in formation among study participants. The adherent to treatment approach will exclude participants from the experimental group who did not engage with app throughout the study. We further plan to quantify temporal changes within participants assigned to the experimental group.

To evaluate changes in sleep behavior, we will examine the distribution of the data to determine if it meets assumptions for parametric tests. Assuming randomization has balanced known and unknown confounders and we observe a normal distribution, we will compare sleep duration between the groups using a t-test. We will use a rank sum test if the distribution is not normal. The distribution of the data will be examined, and transformations will be performed as necessary for valid comparisons. If potential confounders are not balanced between groups, we plan to use regression models with an appropriate link function and distribution to test all hypotheses. The number of healthy sleep changes at the end of the study will be compared between the groups using a rank sum test.

To evaluate changes in sleep quality, we will compare the proportion of participants with poor sleep quality according to the PSQI in the experimental group versus the waitlist control group using a chi-2 test. These continuous measures will be compared using a t-test. To examine changes in mood, alertness, and energy we will compare these ordinal variables using Wilcoxon rank sum tests between the experimental and waitlist control arms. We will estimate changes in absences and productivity using reported salary information and compare the cumulative cost of each using a t-test. We will estimate the incidence of crashes, near-crashes, and injuries which will then be compared using Poisson regression. Finally, we will compare cumulative healthcare costs over the study using t-tests.

## Discussion

Nearly 70% of Americans report routinely curtailing their sleep each month,^1^ and sleep disorders (affecting approximately 50–70 million)^3^ are prevalent among US adults. Although strong evidence demonstrates that worksite-based sleep health education and sleep disorders screening and treatment can improve employee sleep and health, as well as workplace safety, productivity, and healthcare costs,^6,7,10,11^ such programs remain a minority of wellness initiatives reported by employers in the US.^5^

Digital and mobile technologies, such as smartphone devices and wearable sleep and activity monitors, provide several benefits from the standpoint of workplace wellness. First, digital and mobile technologies often feature sensors that can track activities, such as sleep and exercise, and enable personalized messages based on the user and their unique needs.^14,15^ Second, digital and smartphone technologies have the potential to reach employees outside the workday with health promotional messaging. Finally, digital and mobile tools may also be a cost-effective solution when viewed in comparison to high cost experts or coaches.

In this manuscript we describe an operational trial that aims to examine the sleep training smartphone app “dayzz.” The dayzz app offers a mHealth approach with personalized sleep training to improve sleep and address sleep issues. We will monitor participants for up to 9 months to examine the feasibility, acceptability, effectiveness and durability of the dayzz app for improving employee sleep and workplace outcomes in a large employer in the US Northeast. We intend to disseminate the findings by publishing the results in a peer-reviewed journal.

This clinical trial was originally scheduled to begin recruitment with a series of in-person events in March 2020. Our planned recruitment kickoff coincided with the date that the WHO declared COVID-19 a worldwide pandemic.21 After this declaration from the WHO, our study team, like many other study teams,22 made the difficult decision to pause the planned recruitment efforts and re-imagined all aspects of it to be completely virtual including transitioning the SHAW program from in-person to a video format. After successfully transitioning all study policies and procedures, we launched the trial in August 2020.

This randomized trial tests whether the dayzz app is effective in improving sleep, health and well-being of employees. Improvements in employee and employer outcomes observed in this trial would offer strong evidence that, despite the stressors of the current pandemic environment, a smartphone-based solution may be effective for improving sleep and workplace outcomes. A smartphone-based approach to improve sleep and sleep disorders has the potential to be a viable strategy for improving employee health, safety, and well-being. Results of this trial will indicate whether dayzz is a feasible, acceptable and effective approach as an employer-based sleep training and sleep disorder care program.

## Data Availability

No datasets have been generated or analyzed. This submission is a protocol paper. The data availability policy is not applicable to this article.

## Contributors’ Statement

LKB, CAC, SFQ, and MDW conceived of the study. LKB, CAC, SFQ, MDW, RR, JSP, and SQ initiated the study design and implementation of study procedures. LKB is the grant holder. MDW provided statistical expertise in clinical trial design and is conducting the primary statistical analysis. MDW, SQ, JSP, LKB, and RR will have access to the final, anonymized dataset. MCZ and LG advised on the use of the Dayzz app, provided Dayzz videos to support recruitment, identified outcomes of interest. All authors contributed to refinement of the study protocol and approved the final manuscript.

